# Intraoperative sodium range affects white matter microstructure in neonatal congenital heart disease

**DOI:** 10.1101/2025.10.16.25338157

**Authors:** Mirthe E.M. van der Meijden, Kim van Loon, Maaike Nijman, Hanna Talacua, Johannes M.P.J. Breur, Joppe Nijman, Nathalie H.P. Claessens, Serena J. Counsell, Manon J.N.L. Benders, Alexandra F. Bonthrone

**Affiliations:** Department of Neonatology, Wilhelmina Children’s Hospital Utrecht, University Medical Center Utrecht, The Netherlands; Congenital Heart Center Utrecht, Wilhelmina Children’s Hospital, University Medical Center Utrecht, The Netherlands; Research Department of Early Life Imaging, Centre for the Developing Brain, School of Biomedical Engineering and Imaging Sciences, King’s College London, United Kingdom; Department of Pediatric Anesthesiology, Wilhelmina Children’s Hospital Utrecht, University Medical Center Utrecht, The Netherlands; Brain Center Rudolph Magnus, University Medical Center Utrecht, The Netherlands

**Keywords:** congenital heart disease, neonatal cardiac surgery, cardiopulmonary bypass, diffusion weighted imaging, diffusion tensor imaging, sodium course

## Abstract

**Objective:** Intraoperative sodium changes and new postoperative white matter injury are prevalent in neonates with congenital heart disease (CHD) who undergo cardiopulmonary bypass surgery. Rapid sodium correction in hyponatremia is associated with white matter injury in adults. This study examined the potential effects of a rapid intraoperative sodium increase on white matter microstructure in neonates with CHD.

**Methods:** 83 neonates with CHD underwent postoperative diffusion weighted magnetic resonance imaging within three weeks of cardiopulmonary bypass surgery as part of routine clinical practice. Mean diffusivity, radial diffusivity, and axial diffusivity were calculated using Diffusion Tensor Imaging. Voxel-wise associations within the core of white matter tracts were assessed using tract-based spatial statistics. Serum sodium measurements were extracted from clinical notes. Maximum intraoperative sodium range was defined as the difference between the minimum and maximum serum sodium observations during surgery. The rate of sodium change was calculated as this range divided by the time interval in hours.

**Results:** A larger maximum intraoperative sodium range was associated with lower axial diffusivity values in the right centrum semiovale, bilateral precentral white matter, right inferior longitudinal fasciculus, and right optic radiation (p<0.05, family wise error rate corrected). There were no other associations between diffusivities and sodium range or rate of change.

**Conclusions:** A larger maximum intraoperative sodium range was associated with reduced axial diffusivity, possibly indicating axonal injury in neonates with CHD after cardiopulmonary bypass surgery. These findings underscore the importance of limiting perioperative osmotic stress to optimize white matter microstructural development.

**Central picture:** White matter regions affected by intraoperative sodium range.

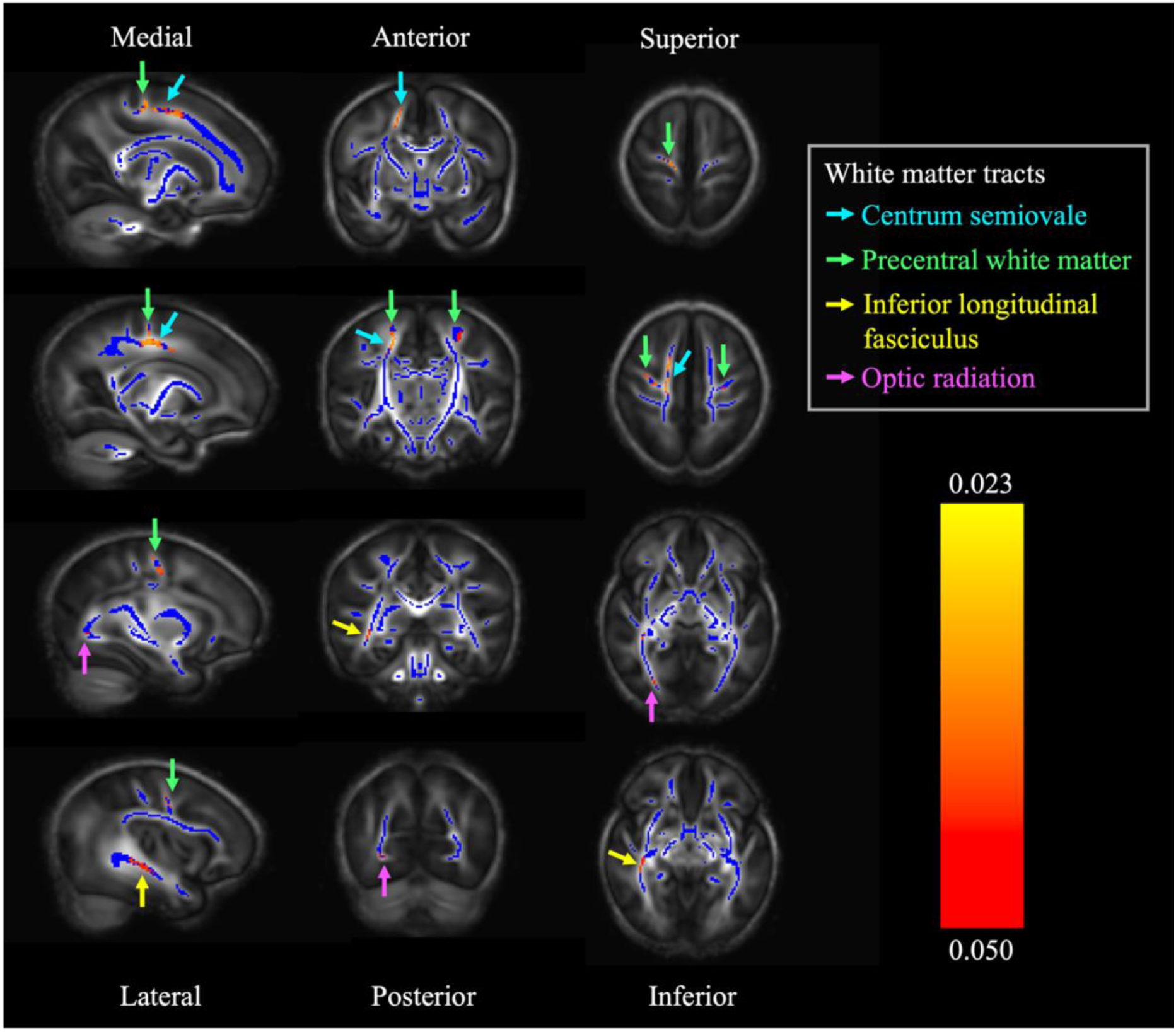

**Central message:** Intraoperative serum sodium changes are associated with white matter microstructural alterations in neonates with congenital heart disease undergoing cardiac surgery with cardiopulmonary bypass.

**Perspective statement:** Rapid sodium changes are common in neonatal congenital heart disease surgery and are associated with white matter injury in other clinical populations. This study shows altered white matter microstructure in neonates with larger intraoperative sodium ranges, emphasizing the importance of limiting perioperative osmotic stress to optimise white matter microstructural development in this population.

## Introduction

Congenital heart disease (CHD) is the most prevalent congenital anomaly, affecting the development of cardiac structures and great vessels.^1^ Although advances in neonatal cardiac surgery and critical care have increased survival rates throughout the past decades, long-term neurodevelopmental impairment remains prevalent.^2^

Altered brain development in CHD originates in utero where altered circulation provides suboptimal cerebral substrate delivery^3^, resulting in smaller brain volumes^3^ and reduced cortical folding^4^ that persists into the neonatal period.^5,6^ Diffusion-weighted magnetic resonance imaging (dMRI) studies have shown pre- and postoperative white matter (WM) microstructural alterations in neonates with CHD, including high mean diffusivity (MD) and low fractional anisotropy (FA) compared to heart-healthy neonates.^7–9^ White matter injury (WMI) is prevalent before and after surgery.^10^ Surgical factors such as neonatal cardiopulmonary bypass (CPB) further increase the risk for new postoperative WMI^10^ and altered brain development in early childhood.^11^ However, risk factors for postoperative brain changes are multifactorial and not well understood, and studies examining the impact of other potential perioperative risk factors are required.

During CPB surgery, administration of hyperosmolar priming solutions induces a rapid sodium increase when baseline is low, which increases the risk for osmotic demyelination syndrome (ODS) in adults.^12^ ODS is a phenomenon characterised by WMI and altered WM microstructure related to a rapid correction of hyponatremia.^13,14^ On dMRI, adults with ODS show a decrease of apparent diffusion coefficient (ADC), a comparable measure to MD, within the first week, followed by an increase to normal levels around one month after the rapid sodium increase.^13,14^ A rapid increase of serum osmolality is hypothesised to induce transcellular water shifts, glial cell shrinkage, and compensatory upregulation of ion transporters and osmolyte synthesis, a metabolically taxing process that depletes adenosine triphosphate.^15^ This osmotic stress disrupts tight junctions and the blood-brain barrier, allowing inflammatory mediators to enter the central nervous system and initiate oligodendrocyte apoptosis and demyelination.

Neonates^16–19^ and children^16–18,20^ with CHD show a rapid sodium increase throughout surgery, possibly increasing the vulnerability to new postoperative WMI^10^ and WM microstructural alterations.^7–9^ In our previous study quantifying the perioperative sodium course in neonates with CHD, the median maximum intraoperative sodium range was 7.0 *mmol/L*.^19^ This approaches the daily threshold of 8.0 *mmol/L* for a safe sodium correction in adults.^21^ Currently, there is no agreed threshold in neonates. We did not report an association between intraoperative sodium changes and new postoperative WMI on qualitative imaging.^19^ However, it is plausible that osmotic stress may result in altered WM microstructure rather than overt WMI visible on structural MRI.

This study aimed to assess the relationship between a rapid sodium increase during cardiac surgery with CPB and WM microstructural alterations in neonates with CHD using Diffusion Tensor Imaging (DTI). We hypothesised reduced postoperative diffusion across the WM in neonates with larger intraoperative sodium changes, reflected by decreased MD, radial diffusivity (RD), and axial diffusivity (AD).

## Methods

### Ethical approval and consent

The Dutch institutional review board approved the study protocol on 17/02/2016 (MREC:16-093). The study adheres to the moral and ethical principles described in the Declaration of Helsinki and good clinical practice.^22^ Informed written consent was provided by parents regarding the usage of imaging and clinical data for research purposes.

### Study population and design

From the original cohort described by van der Meijden et al.^19^, neonates were eligible for inclusion if they were born after 36.0 gestational weeks and underwent dMRI within three weeks after CPB surgery. Of 158 neonates in the original cohort, 128 neonates who underwent surgery within the first six weeks of life between February 2016 and September 2021 at the Wilhelmina Children’s Hospital Utrecht, The Netherlands, were eligible for inclusion in the study.

### Sodium data collection

Sodium data collection was conducted according to the methods described by van der Meijden et al.^19^ Briefly, per clinical protocol, serum sodium levels of neonates were obtained throughout their hospital stay, primarily by point-of-care testing (Abbott iSTAT). The maximum intraoperative sodium range was calculated per neonate by subtracting the lowest from the highest serum sodium level during surgery. This measure reflects the effects of both the exposure to high sodium fluids (e.g., CPB-priming solution, blood products, and intravenous fluids) and low-sodium cardioplegia (Custodiol-Köhler solution). The time difference in hours between the minimum and maximum intraoperative sodium measure was computed. The rate of the maximum intraoperative sodium change was calculated by dividing the maximum intraoperative sodium range by the time difference. Clinical data were extracted from electronic medical records.

### MRI acquisition

Scanning procedures have previously been reported by Claessens et al.^23^ Neonates were placed in a vacuum mattress and received dual-layer hearing protection before scanning (adhesive ear covers and over-ear muffs). Heart rate, saturation, and breathing were continuously monitored. Neonates were scanned in natural sleep where possible or sedated with oral chloral hydrate before scanning when necessary (50-60 *mg/kg*). MRI was acquired as part of routine clinical care. Axial dMRI (TR 6500 *ms*; TE 66 *ms*; flip angle 90°; slice thickness 2 *mm*; acquired voxel size 2.25 x 2.25 x 3.00 *mm*; reconstructed voxel size 2.00 x 2.00 x 2.00 *mm*, diffusion gradients 45 directions, one *b* = 0 *s/mm*^2^ image, *b* value = 800 *s/mm*^2^) was acquired on a Philips 3 Tesla system (Philips Medical Systems, Best, Netherlands), using a 32-channel head coil.

### dMRI preprocessing

An overview of participant exclusions is provided in Figure 1. In preparation of dMRI preprocessing, images were visually quality checked for major motion artefacts.

**Figure 1.**
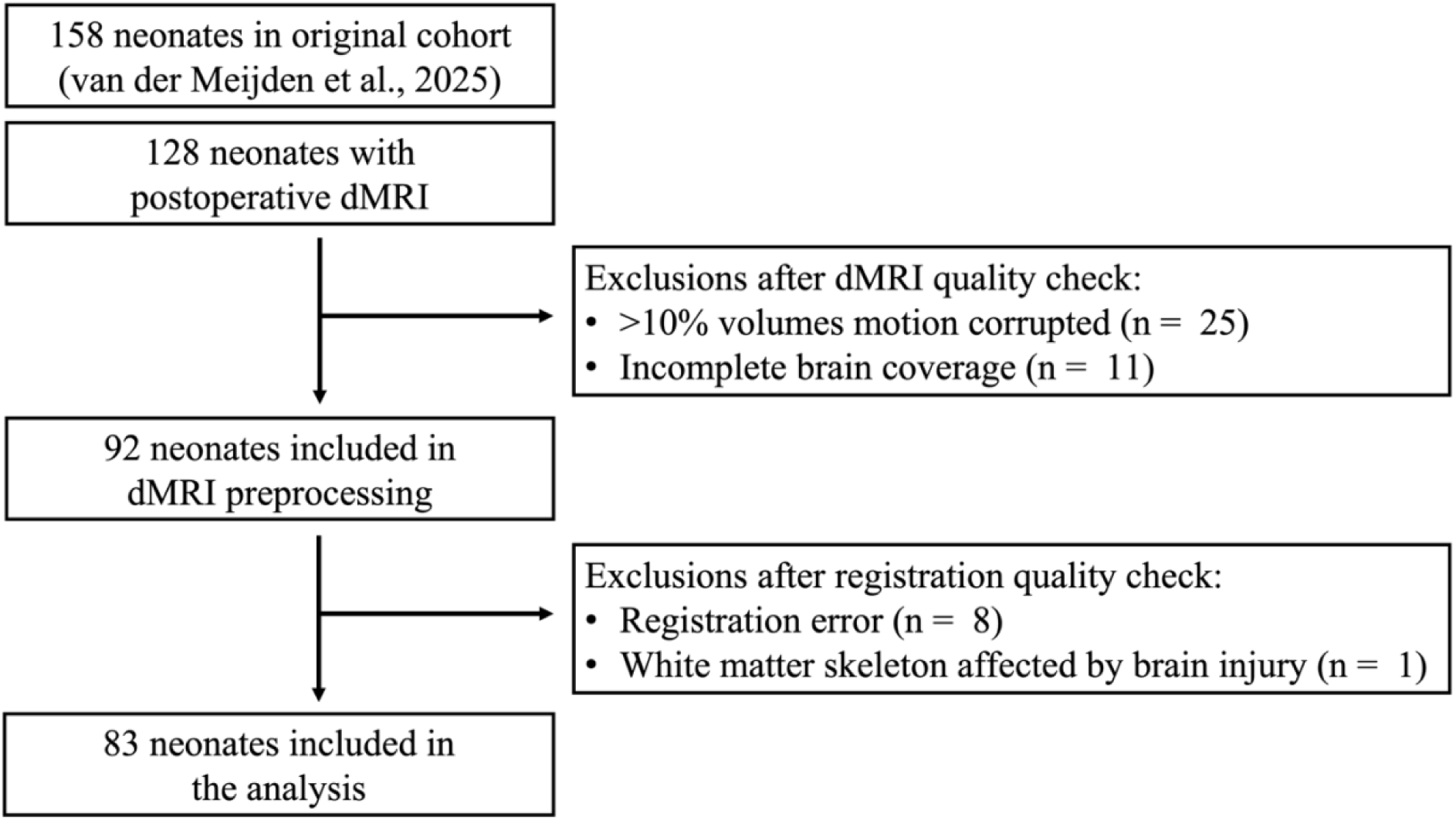
Diagram summarizing exclusions. *Abbreviations*: dMRI, diffusion-weighted Magnetic Resonance Imaging

DMRI images underwent denoising^24,25^ and Gibbs de-ringing^26^ using the MRtrix3 software package (http://www.mrtrix.org).^27^ Images underwent eddy current distortion and motion correction with outlier rejection and replacement using Eddy from the FSL software package (https://fsl.fmrib.ox.ac.uk/fsl/fslwiki).^28^ A quality check report regarding Eddy corrections was generated using Eddy QUality Assessment for DMRI (QUAD). The mean square residuals plot was used to identify volumes for which motion correction was inadequate. These volumes were visually inspected and excluded when major motion artefacts remained apparent. One or more volumes were removed from 51 (55.4%) datasets (median 2, range 1-4 volumes).

Neonatal data processing was undertaken using previously published methods.^29^ A diffusion tensor model was fit per voxel using the dtifit tool in FSL (https://fsl.fmrib.ox.ac.uk/fsl/fslwiki).^30^ The outputs of dtifit were converted to tensor images compatible with DTI-ToolKit (DTI-TK v2.3.3 https://dti-tk.sourceforge.net/pmwiki/pmwiki.php). Registration of dMRI tensor images was undertaken using DTI-TK, a validated tool for accurate registration of neonatal dMRI.^31^ First, an initial population-specific template was created using dMRI datasets of five neonates that were visually evaluated to be of high-quality and adequate anatomical alignment. Subsequently, rigid-body, affine, and deformable registration were used to align the dMRI data. A high-resolution study specific template was computed by averaging the aligned dMRI images.

Commonly used DTI metrics include MD, FA, RD, and AD, corresponding to the average magnitude of water molecular diffusion, the degree of directionality of water molecular motion, and magnitude of water molecular motion perpendicular and parallel to the principal vector per voxel, respectively.^32^ The template was used to extract the WM skeleton, comprising the locally maximal FA values reflective of the centre of the WM tracts of the population. MD, RD, and AD maps were computed per neonate and projected onto the skeleton, enabling voxel-wise analyses across neonates. FA was not assessed, as this metric was not expected to change within the short observational period.^33^

### Statistical analysis

Analyses were carried out using FSL (v6.0 https://fsl.fmrib.ox.ac.uk/fsl/fslwiki) and RStudio (v1.3.1093).^34^ Normality of continuous variables was evaluated using the Shapiro-wilk test. Normally distributed variables are presented as mean ± *SD*, non-normally distributed variables as median (*IQR*), and categorical variables as number (%). In order to assess voxel-wise relationships between maximum sodium range and rate of change, and postoperative MD, RD, and AD, permutation testing was carried out using the randomise tool in FSL (v6.0 https://fsl.fmrib.ox.ac.uk/fsl/fslwiki), using 10000 permutations and Threshold-

Free Cluster Enhancement^35^, accounting for sex, gestational age at birth, postnatal age at surgery, postmenstrual age at scan, interval between surgery and scan, presence of single ventricle physiology, and CPB duration. To improve model fit, all numerical variables were standardised by subtracting the mean from the observed value and dividing by the standard deviation.

## Results

The final cohort consisted of 83 neonates with a total of 2,533 validated plasma sodium observations. Baseline characteristics are summarized in Table 1.

**Table 1.**
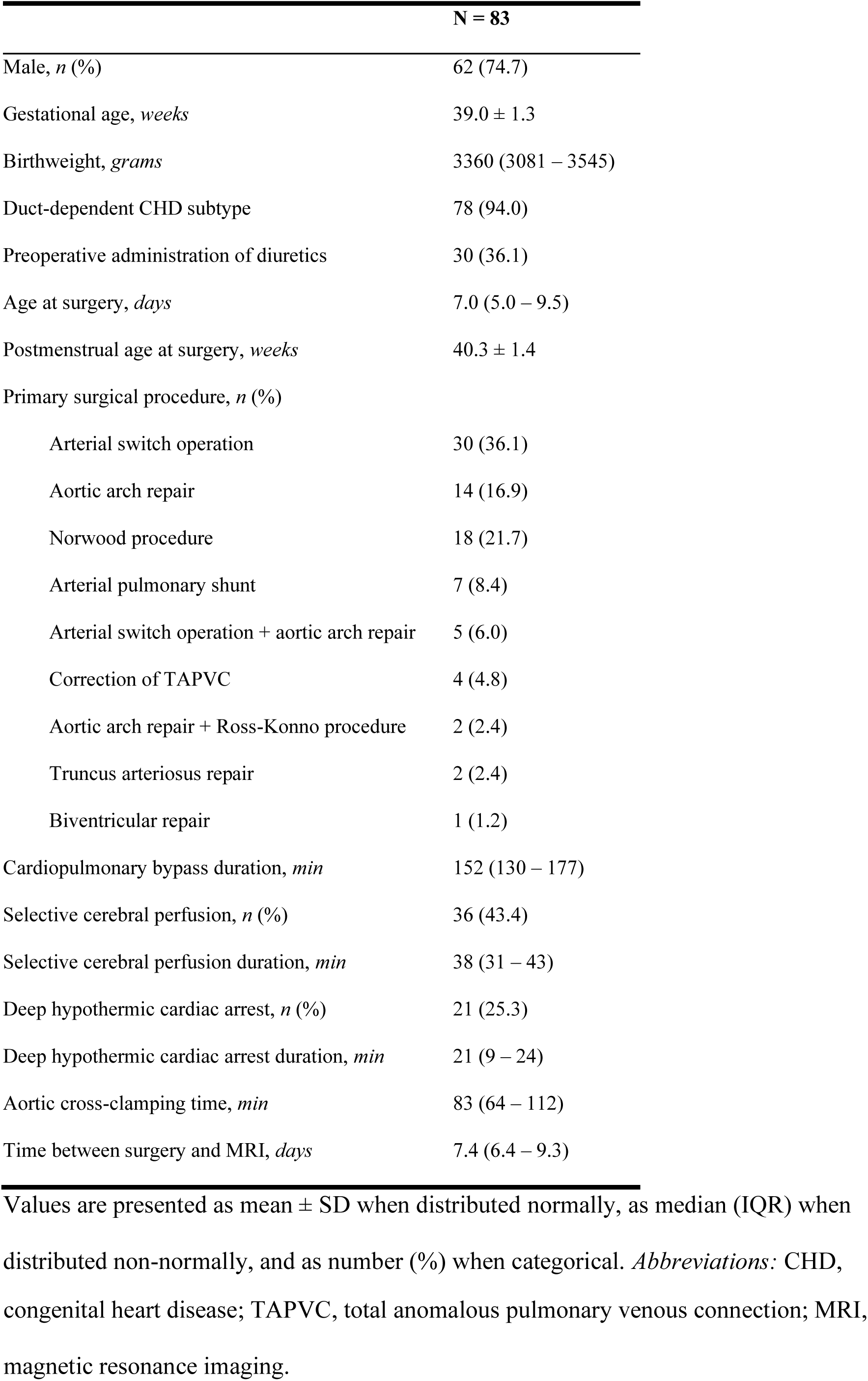
Baseline characteristics.

### Intraoperative sodium characteristics

The intraoperative sodium course is summarized in Table 2. The mean maximum sodium range was 7.7 *mmol/L* (*SD* 3.2), within a median surgery duration of 5.6 hours (*IQR* 4.8–6.1). The advised daily limit of safe sodium correction of 8 *mmol/L* for adults^21^ was exceeded by 37.3% of neonates. The corresponding rate of safe sodium correction for the median surgery duration was 1.9 *mmol/L* (calculated as 8/24*5.6), which was exceeded by 97.6% of neonates. The median rate of the maximum intraoperative sodium increase was 2.86 *mmol/h* (*IQR* −5.3–5.0), which was almost 9 times larger than the hourly rate of safe sodium correction of 0.33 *mmol/L/h* in adults (calculated as 8/24).

**Table 2.** Intraoperative sodium course.

|  | N | Average | Min-Max |
| --- | --- | --- | --- |
| Sodium level, *mmol* | 83 | 139.0 (135.0 – 141.0) | 125.0 – 152.0 |
| Pre-CPB sodium, *mmol* | 67 | 137.4 ± 3.5 | 128.0 – 144.0 |
| CPB priming solution, *mmol* | 76 | 167.8 ± 3.4 | 158.0 - 178.0 |
| Max. sodium range, *mmol* | 83 | 7.7 ± 3.2 | 1.0 – 16.0 |
| Max. sodium range interval, *h* | 83 | 0.67 (-0.80 – 1.81) | -3.88 – 4.30 |
| Max. sodium increase rate, *mmol/h* | 83 | 2.86 (-5.29 – 5.0) | -50.0 – 34.29 |

Values are presented as mean ± SD when distributed normally, as median (IQR) when distributed non-normally, and as number (%) when categorical. *Abbreviations:* CPB, cardiopulmonary bypass

### Voxel-wise analyses

Voxel-wise analysis of postoperative AD revealed that a higher maximum intraoperative sodium range was associated with lower AD values in the right centrum semiovale, precentral WM (right>left), right inferior longitudinal fasciculus, and right optic radiation (p<0.05) (Figure 2). The relationship between mean AD within the significant voxels of the centrum semiovale and the maximum intraoperative sodium range adjusted for the covariates is presented in Figure 3. MD and RD were not associated with maximum intraoperative sodium range. No relationship was evident between postoperative MD, AD, or RD and the rate of the maximum intraoperative sodium increase.

**Figure 2.**
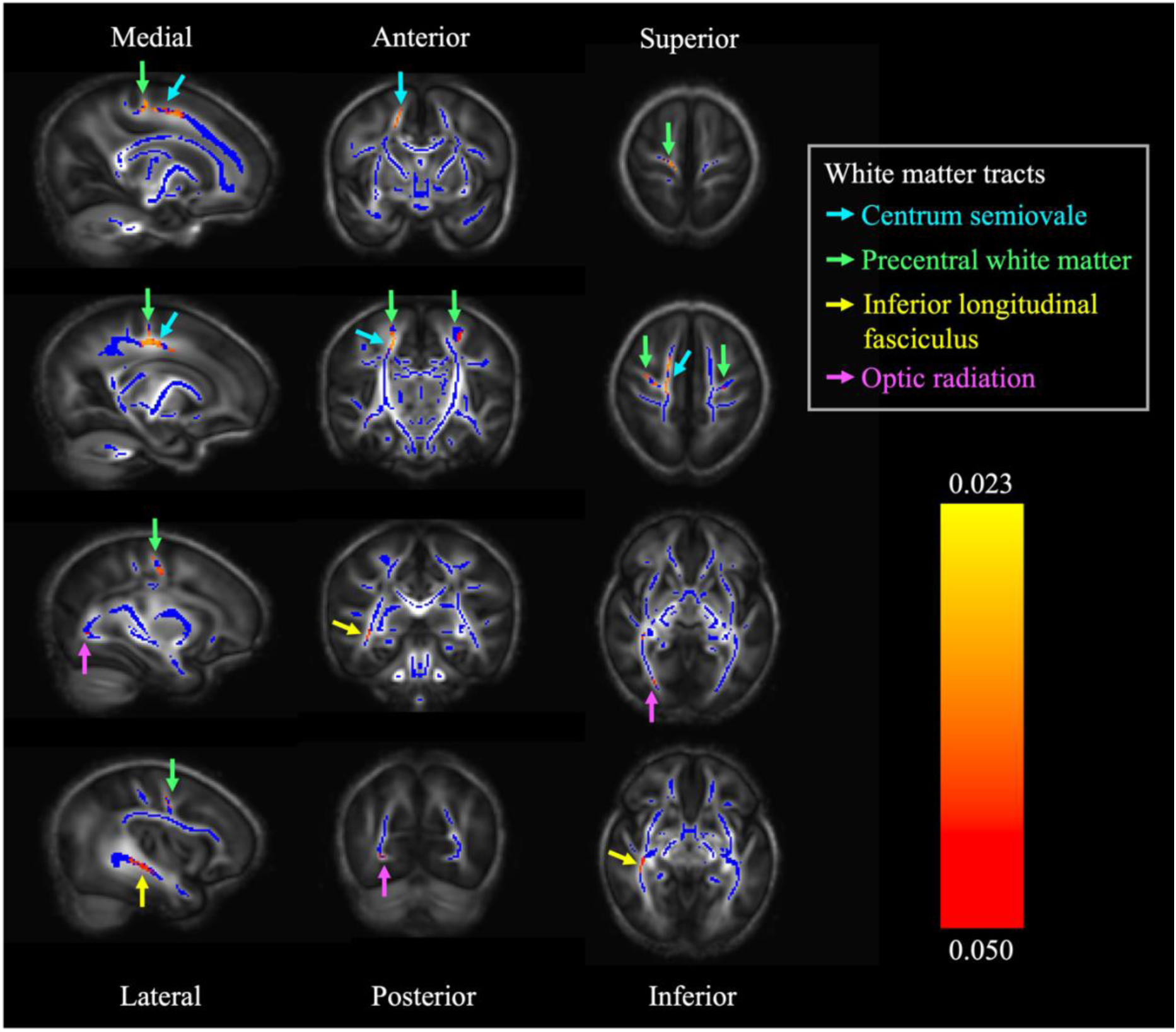
White matter regions that show a significant negative relationship between maximum intraoperative sodium range and postoperative AD (red/orange/yellow, p<0.05), overlaid on FA skeleton (blue) and mean FA map (grey). All sagittal images are selected from the right hemisphere.

**Figure 3.**
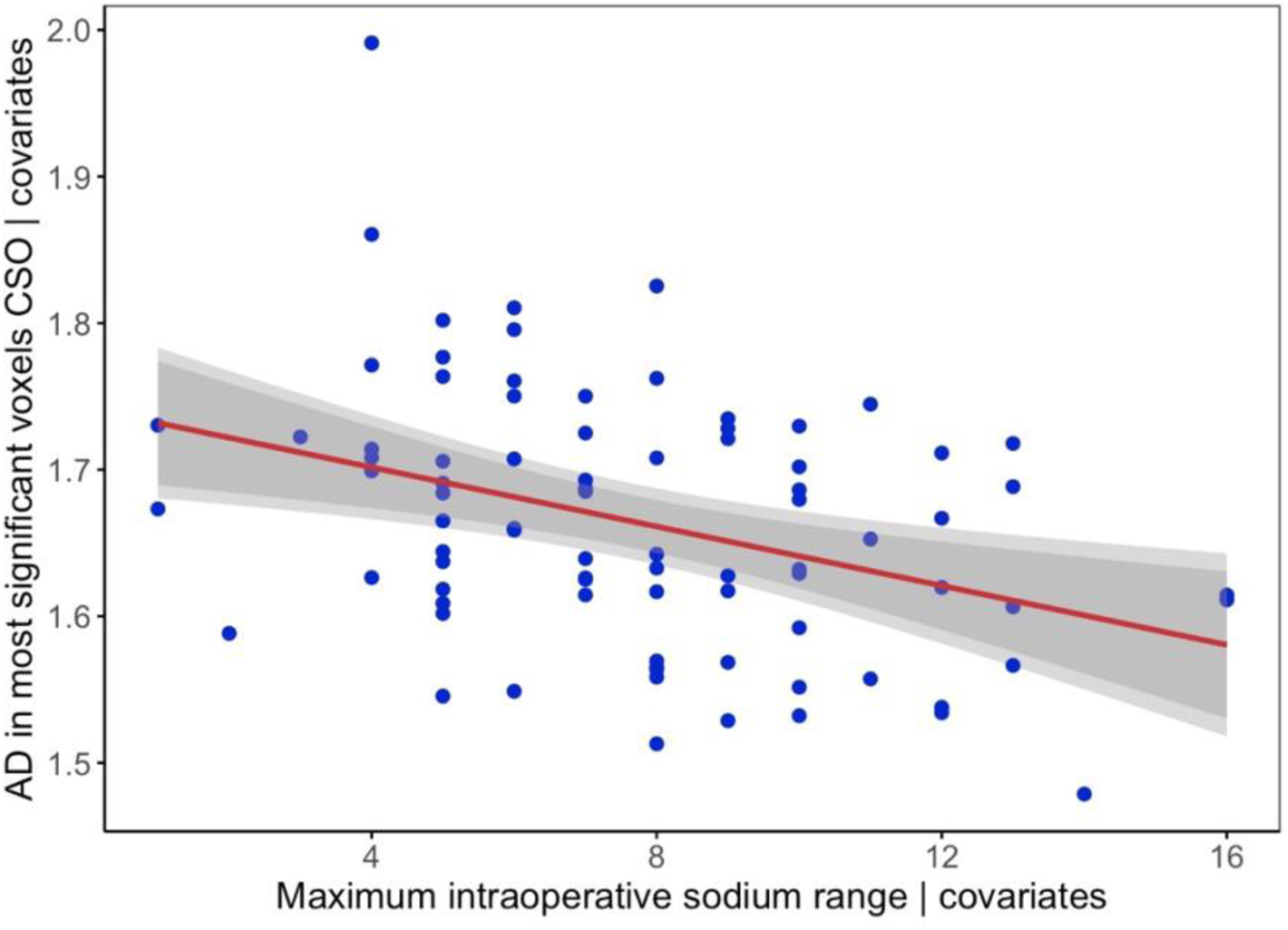
Scatterplot showing the relationship between mean AD in the significant voxels within the centrum semiovale and maximum intraoperative sodium range adjusted for the covariates. *Abbreviations*: CSO, centrum semiovale; AD, axial diffusivity

## Discussion

To our knowledge, this is the first study examining the effects of serum sodium rise during cardiac surgery on WM microstructure in neonates with CHD. Concordant with our hypothesis, we report that larger intraoperative sodium ranges were associated with decreased AD in the right centrum semiovale, bilateral precentral WM, right inferior longitudinal fasciculus, and right optic radiation. The rate of the maximum intraoperative sodium change was not associated with MD, AD, or RD.

The observation of reduced AD in WM of neonates with a larger intraoperative sodium range is partially aligned with adult ODS case reports, showing reduced ADC, a measure of average diffusivity, approximately one week after a rapid sodium increase.^13,14^ However, to our knowledge, adult ODS studies have not assessed changes in AD and RD. The observation of reduced AD, but not MD and RD, may therefore be better understood by considering diffusion changes in other neonatal conditions in which oligodendrocyte injury is prevalent, such as hypoxic-ischemic encephalopathy. Neonates with hypoxic-ischemic encephalopathy show an initial decrease of MD after ischemia^36^, similar to the decrease of ADC observed in adult ODS. Additionally, a neonatal mouse model of retinal ischemia has provided a more detailed pattern of diffusivity changes, indicating initial reduced AD and MD within the first three days after ischemia, followed by increased RD and MD.^37^ These diffusivity changes are reflective of axonal injury and demyelination, respectively, corroborated by histological reports. It is possible that our observations reflect the phase of axonal injury where AD remains low, but RD and MD have started to normalize. However, adults with ODS show normalization of ADC approximately one month after the rapid sodium increase^13,14^, whereas normalization in neonates with hypoxic-ischemic encephalopathy occurs between 6 and 12 days after ischemia^36^. We are unaware of any studies assessing the relationship between sodium changes, diffusivities and histological changes in the neonatal brain. Neonates in the current cohort were scanned median 7.4 days after surgery, hence the observed decreased AD could be more aligned with the slower time course of diffusivity changes in adult ODS. However, adult ODS is typically observed after rapid correction of hyponatremia, and we previously reported that only 31% of neonates are hyponatremic in the 72h before CPB surgery.^19^ More research is needed to clarify the exact time course and underlying mechanisms of WM microstructure alterations in relation to intraoperative sodium changes in neonates with CHD. MRI at 3 months post-surgery could provide valuable insight into the time course of WM microstructure alterations in response to intraoperative sodium change.

The reasons underlying the regional specificity of the effect of maximum intraoperative sodium range in our study are unclear. It is possible that some WM regions are more susceptible to osmotic stress than others. One study examining brain injury in neonatal hypernatremic dehydration, a condition where water is transported from the body at a faster rate than sodium, reported cytotoxic edema in the centrum semiovale.^38^ Although the underlying conditions differ, this finding supports the current finding that the centrum semiovale may be particularly sensitive to osmotic fluctuations. However, there have been no previous studies linking the precentral WM, inferior longitudinal fasciculus, or optic radiation to sodium changes. Further investigation is needed to understand why these regions may be particularly vulnerable to osmotic stress.

Current findings expand on our previous work^19^, underscoring the importance of close monitoring and minimization of osmotic stress during CPB surgery in neonates with CHD. To limit the range of intraoperative sodium observations and reduce fluid shifts, low- and high-sodium fluids should be administered with caution. The perioperative protocol at Wilhelmina Children’s Hospital has been changed accordingly, by diluting vasoactive medication in glucose 10% to reduce sodium load.

Interpretation of our results is complicated by the complex multifactorial origin of perioperative brain alterations in CHD, where it is difficult to isolate the effects of osmotic stress maximum intraoperative sodium range on WM microstructure from other surgical and clinical factors. Hemodynamically unstable neonates receive more intraoperative fluids, possibly increasing the range of sodium values and potentiating fluid shifts. Moreover, indicators of hemodynamic instability such as low cardiac output, are associated with increased risk of brain injury.^24^ Maximum sodium range could be a marker of hemodynamic instability rather than a causal factor for alterations in WM. Additionally, we did not account for all surgical variables in the TBSS model, as diagnosis, surgical procedure, and perioperative factors are highly correlated. We included presence of single ventricle physiology and duration of cardiopulmonary pass, due to their established association with new WMI.^10,39^ Lastly, not all neonates from the original cohort^19^ had dMRI available.

Clinical instability was the primary reason MRI was not obtained within three weeks after surgery, this cohort therefore likely reflects the effects of sodium in more clinically stable neonates with CHD.

In conclusion, the current study suggests a large intraoperative sodium range is associated with altered WM microstructure in the centrum semiovale, precentral WM, inferior longitudinal fasciculus, and optic radiation. This finding may have important implications for perioperative sodium regulation protocols to support WM microstructure development in neonates with CHD undergoing CPB surgery.

## Data Availability

The data used in this study are available from the corresponding author upon reasonable request. Some types of data sharing may be restricted by the informed consent statement of individual patients.

## Glossary

CHD: congenital heart disease
WM: white matter
WMI: white matter injury
dMRI: diffusion magnetic resonance imaging
CPB: cardiopulmonary bypass
ODS: osmotic demyelination syndrome
DTI: diffusion tensor imaging
ADC: apparent diffusion coefficient
MD: mean diffusivity
RD: radial diffusivity
AD: axial diffusivity
FA: fractional anisotropy

## Disclosure

Authors report no conflicts of interest regarding the content of this work.

## Funding

Regular funding was provided to Dutch universities to advance medical research. MEMVDM, SJC, and AFB are supported by the UKRI Medical Research Council [MR/V002465/1].

## Acknowledgements

We thank the families for participating in this study. We also thank the Congenital Heart Disease Life Span study group of the Wilhelmina Children’s Hospital, including the Departments of Neonatology, Pediatric Intensive Care, Pediatric Cardiology, Pediatric Cardiothoracic Surgery, Pediatric Anesthesiology, and Radiology.

## Notes

### Competing Interest Statement

The authors have declared no competing interest.

### Funding Statement

This study was funded by regular funding provided to Dutch universities to advance medical research. Mirthe E.M. van der Meijden, Serena J. Counsell, and Alexandra F. Bonthrone are supported by the UKRI Medical Research Council [MR/V002465/1].

### Author Declarations

The Institutional Review Board of the Medical Research Ethics Committee gave ethical approval for this work on 17 February 2016 (MREC: 16-093)

